# Deceased-donor kidney transplantation in Brazil, 2014–2024: temporal patterns, regional heterogeneity, and donation-context indicators

**DOI:** 10.64898/2026.06.26.26356660

**Authors:** Márcia Bastos Convento, Fernanda Teixeira Borges

**Affiliations:** Federal University of São Paulo (UNIFESP), Paulista School of Medicine, Department of Medicine, Division of Nephrology, São Paulo, SP, Brazil; Cruzeiro do Sul University, Health Sciences, São Paulo, SP, Brazil

**Keywords:** Kidney transplantation, deceased donor, organ donation, family refusal, regional disparities

## Abstract

**Background:** Deceased-donor kidney transplantation is a component of the Brazilian transplant system and takes place within a deceased-donation environment that includes donor identification, notification, family approach and authorization, organ allocation, and logistics. This study described temporal, macroregional, and federative unit-level variation in deceased-donor kidney transplantation in Brazil from 2014 to 2024, together with hospitalization-based indicators and contextual indicators of the deceased-donation environment.

**Methods:** This nationwide descriptive time-series study used publicly available secondary data from the Hospital Information System of the Unified Health System (SIH/SUS), the Brazilian Transplant Registry (RBT/ABTO), and the Brazilian National Transplant System (SNT). Indicators included the number of transplants, transplant rates per million population, kidney transplant waiting list stock, mean length of hospital stay, in-hospital mortality, potential donor notifications, and family interview and refusal proportions. The three databases were analyzed separately and in parallel, without linkage.

**Results:** The annual number of deceased-donor kidney transplants increased over the study period. The kidney transplant waiting list stock also increased. Mean length of hospital stay decreased, and in-hospital mortality decreased over time. Marked macroregional and federative unit-level heterogeneity was observed in transplant activity, hospitalization-based indicators, and contextual indicators of the deceased-donation environment.

**Conclusions:** Deceased-donor kidney transplantation increased in Brazil between 2014 and 2024, although regional disparities persisted. These findings support monitoring strategies that incorporate contextual indicators alongside measures of transplant activity. Because this was a descriptive study based on secondary data from distinct sources, the findings should not be interpreted as evidence of causal relationships.

## 1. Introduction

Chronic kidney disease (CKD) is a major public health challenge, with high morbidity and mortality, impaired quality of life, and substantial healthcare costs¹. In advanced stages, kidney replacement therapy is required and includes dialysis and kidney transplantation². Among these modalities, kidney transplantation is the preferred treatment for eligible patients because it has been associated with better survival, improved quality of life, and greater long-term cost-effectiveness than maintenance dialysis³.

In Brazil, kidney transplantation plays a central role in the national transplant system and accounts for the largest share of solid-organ transplants performed nationwide^3^. Deceased-donor kidney transplantation is particularly important for expanding access to treatment; however, it requires a complex and highly coordinated set of clinical and organizational components. These include the identification and notification of potential donors, family approach and authorization, donor maintenance, organ allocation, transportation logistics, and transplant sur-gery⁴,^5^. Thus, deceased-donor kidney transplant activity takes place within a broader deceased-donation environment that includes donor availability, organizational features, and operational processes.

Some contextual operational indicators commonly used to characterize the deceased-donation environment, such as potential donor notifications, family interviews, and family refusal, do not refer specifically to kidney donation itself, but rather to the broader deceased organ donation system. These indicators may therefore be interpreted as contextual markers of the deceased-donation environment in which deceased-donor kidney transplantation takes place, rather than as direct kidney-specific measures.

In addition, regional differences in hospital infrastructure, availability of specialized teams, organization of transplant coordination systems, and geographic and logistical conditions may be relevant contextual features when interpreting variation in transplant activity and contextual indicators of the deceased-donation environment across the country^6^˒^7^.

A nationwide assessment of deceased-donor kidney transplantation in Brazil is relevant because it can describe temporal and territorial variation in transplant activity while situating this activity within the broader deceased-donation environment. In a large and heterogeneous country, such a description may help identify persistent territorial asymmetries that are not fully captured by national totals alone. Therefore, this study aimed to describe temporal and territorial variation in deceased-donor kidney transplantation in Brazil from 2014 to 2024 and to contextualize this variation using hospitalization-based indicators and selected deceased-donation contextual indicators.

## 2. Methods

This nationwide descriptive time-series study used publicly available aggregated secondary data from Brazil for the period 2014–2024. Data on the national kidney transplant waiting list stock were obtained from the Brazilian National Transplant System (SNT)^8^ historical report and correspond to the annual kidney transplant technical registry, defined in the source report as the waiting list for active plus semi-actives potential recipient, as reported by the system for the period 2014–2024.

Hospitalization-based indicators were obtained from the Hospital Information System of the Unified Health System (SIH/SUS)^9^, accessed via DATASUS/TabNet on January 10, 2026. Records were retrieved according to the year of hospitalization. Hospital admissions in which deceased-donor kidney transplantation was recorded as the primary procedure were identified using procedure code 0505020092. Annual macroregional indicators extracted from SIH/SUS as reported by the system included the mean length of stay (days) and in-hospital mortality (%). These indicators refer exclusively to hospitalizations recorded in SIH/SUS and should therefore be interpreted within the scope of this administrative database rather than as national totals for all kidney transplant hospitalizations.

Contextual indicators related to the deceased-donation environment were obtained from the Brazilian Transplant Registry (Registro Brasileiro de Transplantes, RBT), published by the Brazilian Organ Transplant Association (ABTO)^10^, and accessed on January 10, 2026. For the period 2014–2024, data were extracted by macroregions and federative units on the number of deceased-donor kidney transplants, deceased-donor kidney transplant rates per million population (pmp), potential donor notifications, family interviews, and family refusals, as reported by RBT/ABTO.

The annual family interview proportion was calculated as (family interviews / potential donor notifications) × 100, and the annual family refusal proportion as (family refusals / family interviews) × 100. When the annual denominator was zero, the corresponding annual proportion was considered non-estimable. For the summarized presentation of federative unit–level indicators over the 2014–2024 period, mean annual transplant rates were calculated as simple arithmetic means of the annual pmp values observed during the study period. In contrast, the summarized family interview and refusal proportions were calculated using aggregated numerators and denominators across the full study period, yielding denominator-weighted pooled estimates rather than simple arithmetic means of annual proportions.

True zero values were retained as 0.00, whereas summarized proportions were considered non-estimable only when the denominator for the full study period was equal to zero. No imputation was performed for missing or non-estimable values. Source-reported values were retained as originally reported to preserve transparency and reproducibility.

Because potential donor notifications, family interviews, and family refusals are not specific to kidney donation alone, these variables were interpreted as contextual markers of the deceased-donation environment rather than as direct kidney-specific measures. The SNT, SIH/SUS, and RBT/ABTO data were analyzed separately and in parallel, without individual-level linkage between databases.

Data were organized and analyzed descriptively using Microsoft Excel 2024. The analysis included absolute values, rates, proportions, relative changes, and temporal variation in selected indicators. No inferential statistical tests or formal temporal trend models were applied because the study objective was descriptive rather than explanatory.

Because this study used aggregated, publicly available secondary data with no individual identification, it was exempt from review by a Research Ethics Committee, in accordance with Resolution No. 510/2016 of the Brazilian National Health Council.

## 3. Results

### 3.1. National temporal patterns in kidney transplant waiting list stock, deceased-donor kidney transplant activity, and hospitalization-based indicators, 2014–2024

As shown in Table 1, the kidney transplant waiting list stock in Brazil increased overall from 24,297 in 2014 to 39,363 in 2024, corresponding to a 62.01% increase over the study period. After a slight fluctuation between 2014 and 2016, the waiting list increased from 28,351 in 2017 to 30,725 in 2019, declined slightly in 2020 to 30,016, and then rose continuously to 31,764 in 2021, 34,807 in 2022, 38,258 in 2023, and reached its highest value in the series in 2024. Over the same period, the annual number of deceased-donor kidney transplants increased from 4,278 to 5,385, corresponding to a 25.88% increase.

**Table 1.** Annual kidney transplant waiting list stock, deceased-donor kidney transplant activity, and hospitalization-based indicators in Brazil, 2014–2024. Data sources: Brazilian National Transplant System (Sistema Nacional de Transplantes, SNT), SIH/SUS, Ministry of Health⁹, and the Brazilian Transplant Registry (RBT/ABTO)¹⁰.

| Data Source | SNT | RBT/ABTO | RBT/ABTO | SIH/SUS | SIH/SUS |
| --- | --- | --- | --- | --- | --- |
| Year | Waiting list stock (n) | Transplants (n) | Transplants (pmp) | Mean length of stay (days) | In-hospital mortality (%) |
| 2014 | 24,297 | 4,278 | 22.3 | 14.2 | 1.93 |
| 2015 | 25,077 | 4,398 | 21.6 | 13.4 | 1.77 |
| 2016 | 24,914 | 4,293 | 21.0 | 12.5 | 1.67 |
| 2017 | 28,351 | 4,808 | 23.3 | 11.7 | 1.89 |
| 2018 | 28,695 | 4,944 | 23.6 | 11.4 | 1.70 |
| 2019 | 30,725 | 5,229 | 25.0 | 11.1 | 1.60 |
| 2020 | 30,016 | 4,386 | 20.8 | 10.8 | 1.27 |
| 2021 | 31,764 | 4,222 | 19.7 | 10.8 | 1.35 |
| 2022 | 34,807 | 4,634 | 21.4 | 10.5 | 1.21 |
| 2023 | 38,258 | 5,285 | 25.6 | 10.6 | 1.52 |
| 2024 | 39,363 | 5,385 | 25.3 | 10.7 | 1.58 |
| Relative change, 2024 vs 2014 (%) | +62.01 | +25.88 | +13.45 | –24.65 | –18.13 |

**Note:** Waiting list stock was obtained from the Brazilian National Transplant System (SNT), annual transplant counts and rates from the Brazilian Transplant Registry (RBT/ABTO), and hospitalization-based indicators from SIH/SUS. Because these indicators were derived from distinct secondary data systems with different scopes and underlying structures, they should not be interpreted as sharing the same denominator, population universe, or case capture. Values are presented as originally reported in the source systems, without recalculation or adjustment by the authors. Relative change (2024 vs 2014) was calculated by the authors from source-reported annual values.

After relative stability between 2014 and 2016, transplant activity increased from 4,808 in 2017 to 5,229 in 2019, declined in 2020 and 2021 to 4,386 and 4,222, respectively, and then recovered to 4,634 in 2022, 5,285 in 2023, and 5,385 in 2024, the highest annual number observed during the study period.

The transplant rate showed a similar temporal pattern, increasing overall from 22.3 pmp in 2014 to 25.3 pmp in 2024, corresponding to a 13.45% increase. After decreasing to 21.0 pmp in 2016, the rate increased to 25.0 pmp in 2019, fell to 20.8 pmp in 2020 and 19.7 pmp in 2021, and rose again to 21.4 pmp in 2022 and 25.6 pmp in 2023, with a slight decline to 25.3 pmp in 2024. In contrast, the mean length of hospital stay decreased steadily from 14.2 days in 2014 to 10.7 days in 2024, corresponding to a 24.65% reduction, with the lowest value recorded in 2022 (10.5 days).

In-hospital mortality also decreased overall, from 1.93% in 2014 to 1.58% in 2024, corresponding to an 18.13% reduction. After declining through most of the study period and reaching its lowest value in 2022 (1.21%), in-hospital mortality increased slightly in 2023 and 2024, although it remained below the values observed at the beginning of the series.

### 3.2. Macroregional hospitalization-based indicators, 2014–2024

Table 2 summarizes hospitalization-based indicators for deceased-donor kidney transplantation across Brazilian macroregions during 2014–2024, using SIH/SUS records. Mean length of hospital stay was lowest in the Southeast (11.10 days), followed by the South (11.40 days) and the Northeast (12.00 days), and highest in the North (15.00 days) and the Midwest (15.20 days). In-hospital mortality was lowest in the Northeast (1.08%), followed by the North (1.37%), the South (1.56%), and the Southeast (1.77%), and highest in the Midwest (2.08%).

**Table 2.** Hospitalization-based indicators for deceased-donor kidney transplantation across Brazilian macroregions, 2014–2024. Data source: SIH/SUS, Ministry of Health⁹.

| Brazilian macroregions (2014–2024) | Mean length of stay (days) | In-hospital mortality (%) |
| --- | --- | --- |
| North | 15.00 | 1.37 |
| Midwest | 15.20 | 2.08 |
| Northeast | 12.00 | 1.08 |
| Southeast | 11.10 | 1.77 |
| South | 11.40 | 1.56 |

**Note:** Values were extracted from the DATASUS/SIH-SUS database and are presented as originally reported in the source system, without recalculation or adjustment by the authors.

### 3.3. Federative unit–level patterns in deceased-donor kidney transplant activity and contextual indicators of the deceased-donation environment, 2014–2024

Given the marked territorial heterogeneity of Brazil and the study objective of describing variation at both macroregional and federative unit levels, graphical displays were organized by macroregion to preserve state-level patterns that could be diluted in national aggregate presentations. Although this approach increased the visual density of the figures, it allowed simultaneous assessment of temporal variation, transplant activity, and contextual indicators of the deceased-donation environment within each macroregion.

In Figure 1A, annual deceased-donor kidney transplant rates in Southern Brazil varied across the three federative units between 2014 and 2024. Rio Grande do Sul presented the highest mean annual transplant rate for 2014–2024 (39.99 pmp), followed by Paraná (37.99 pmp) and Santa Catarina (35.18 pmp) (Figure 1B).

**Figure 1.**
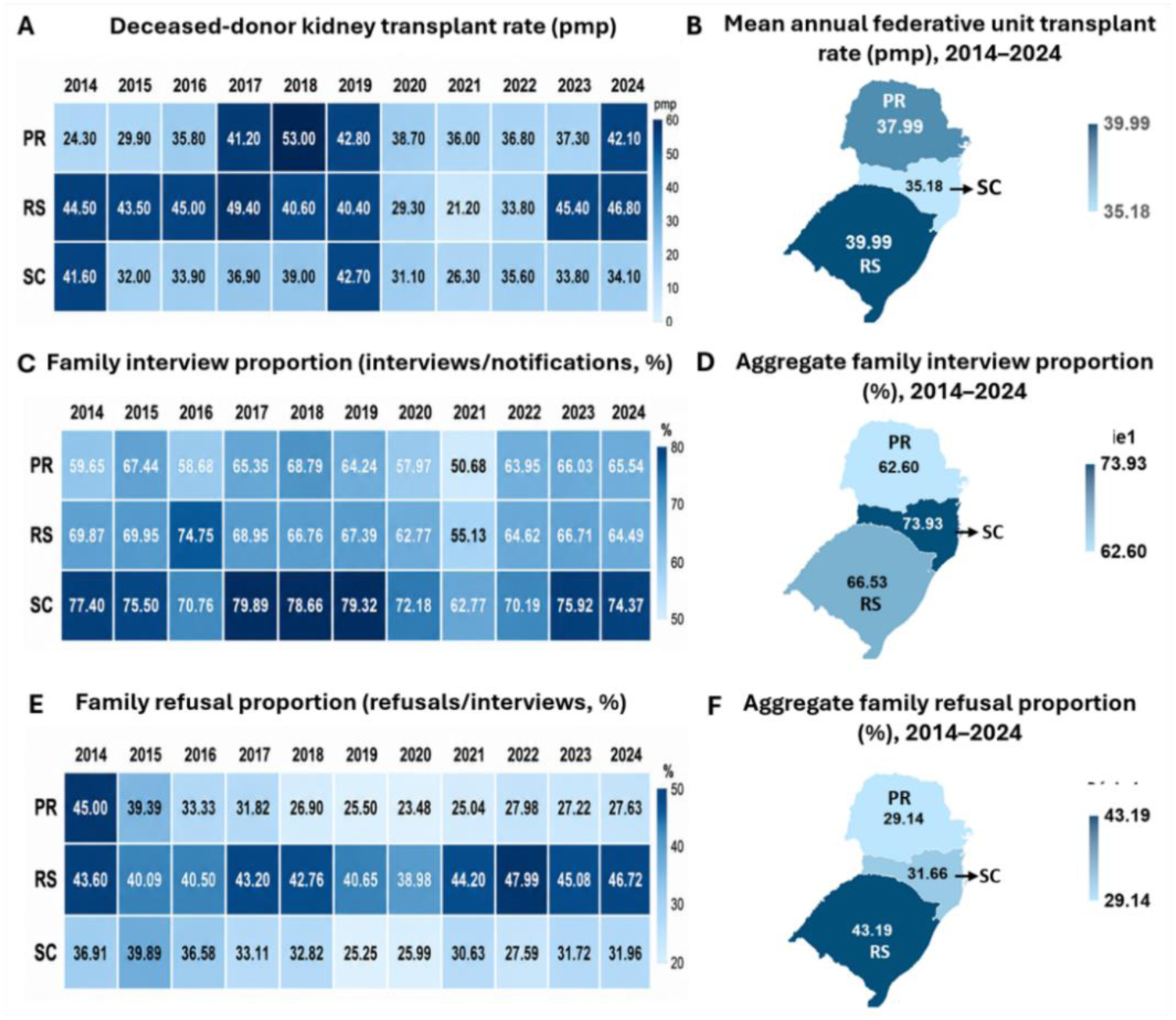
Deceased-donor kidney transplant activity and contextual indicators of the deceased-donation environment in Southern Brazil (Paraná [PR], Rio Grande do Sul [RS], and Santa Catarina [SC]), 2014–2024. The deceased-donor kidney transplant rate per million population is shown as annual federative unit-level values (A) and as a geographic map of the mean annual transplant rate for 2014–2024 (B). Family interview proportion (interviews/notifications, %) is shown as annual federative unit-level values (C) and as a geographic map of the aggregated family interview proportion for 2014–2024, calculated as total interviews divided by total notifications over the study period (D). Family refusal proportion (refusals/interviews, %) is shown as annual federative unit-level values (E) and as a geographic map of the aggregated family refusal proportion for 2014–2024, calculated as total refusals divided by total interviews over the study period (F). **Data source**: the Brazilian Transplant Registry (RBT/ABTO)¹⁰.

In Figure 1C, family interview proportions also varied across the three federative units. Santa Catarina consistently presented the highest values, followed by Rio Grande do Sul and Paraná, with lower values observed in all three states in 2020 and 2021. As summarized in Figure 1D, the aggregated family interview proportion for 2014–2024 was highest in Santa Catarina (73.93%), followed by Rio Grande do Sul (66.53%) and Paraná (62.60%).

In Figure 1E, annual family refusal proportions also varied across the three federative units. Rio Grande do Sul generally presented the highest values, Paraná showed an overall decline over the study period, and Santa Catarina remained below Rio Grande do Sul throughout the series. As shown in Figure 1F, the aggregated family refusal proportion for 2014–2024 was highest in Rio Grande do Sul (43.19%), intermediate in Santa Catarina (31.66%), and lowest in Paraná (29.14%).

In Figure 2A, annual deceased-donor kidney transplant rates in Southeastern Brazil varied across the four federative units between 2014 and 2024. São Paulo consistently presented the highest rates throughout the study period, whereas Espírito Santo showed the lowest values. Rio de Janeiro and Minas Gerais presented intermediate rates, with Minas Gerais approaching the values observed in Rio de Janeiro in the final years of the series.

**Figure 2.**
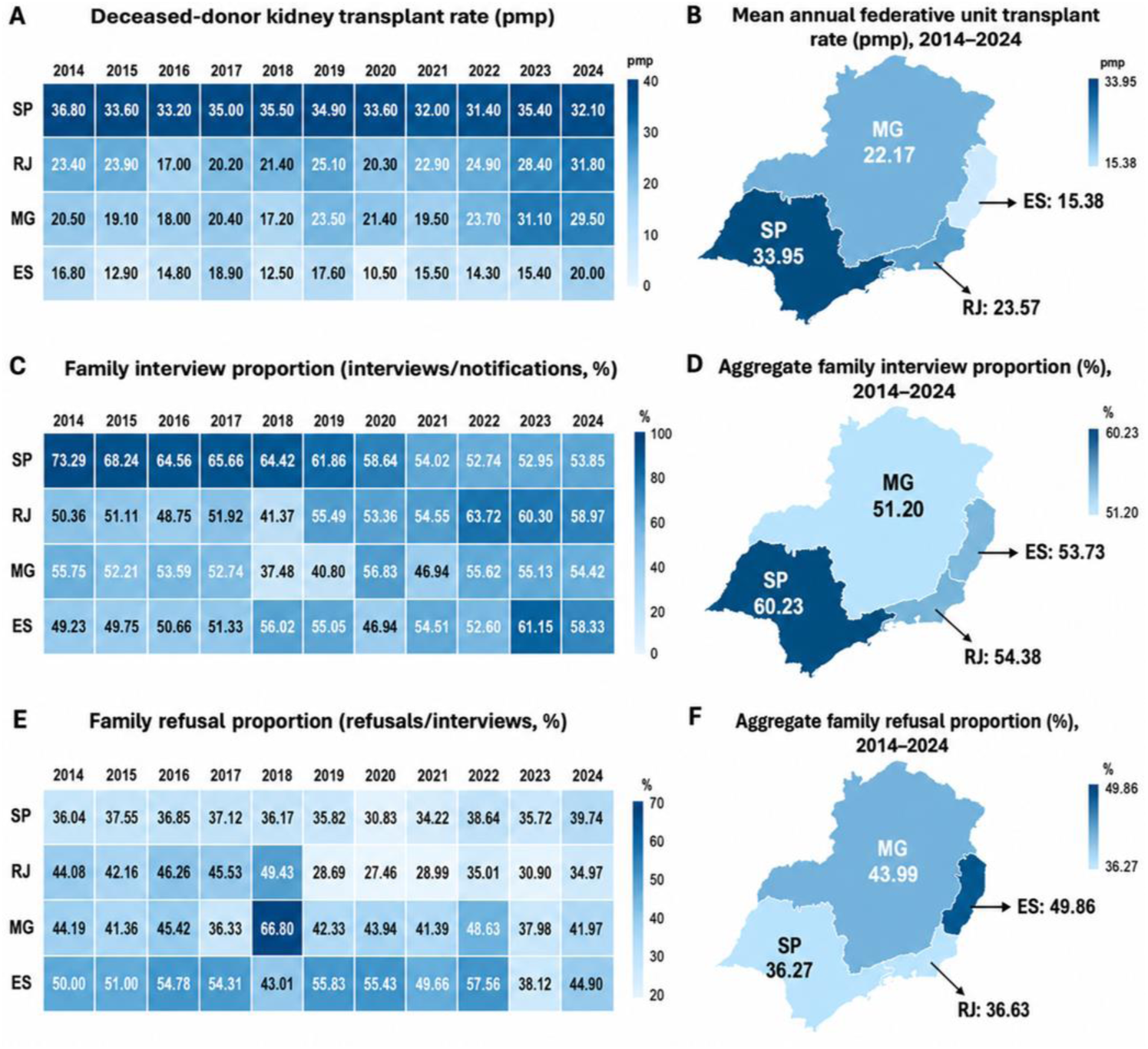
Deceased-donor kidney transplant activity and contextual indicators of the deceased-donation environment in Southeastern Brazil (São Paulo [SP], Rio de Janeiro [RJ], Minas Gerais [MG], and Espírito Santo [ES]), 2014–2024. The deceased-donor kidney transplant rate per million population is shown as annual federative unit-level values (A) and as a geographic map of the mean annual transplant rate for 2014–2024 (B). Family interview proportion (interviews/notifications, %) is shown as annual federative unit-level values (C) and as a geographic map of the aggregated family interview proportion for 2014–2024, calculated as total interviews divided by total notifications over the study period (D). Family refusal proportion (refusals/interviews, %) is shown as annual federative unit-level values (E) and as a geographic map of the aggregated family refusal proportion for 2014–2024, calculated as total refusals divided by total interviews over the study period (F). **Data source**: the Brazilian Transplant Registry (RBT/ABTO)¹⁰.

As shown in Figure 2B, the mean annual transplant rate for 2014–2024 was highest in São Paulo (33.95 pmp), followed by Rio de Janeiro (23.57 pmp), Minas Gerais (22.17 pmp), and Espírito Santo (15.38 pmp).

In Figure 2C, family interview proportions also varied across the federative units. São Paulo showed the highest aggregated proportion for 2014–2024 (60.23%), followed by Rio de Janeiro (54.38%), Espírito Santo (53.73%), and Minas Gerais (51.20%) (Figure 2D).In Figure 2E, annual family refusal proportions were heterogeneous across the four federative units. Espírito Santo generally presented the highest values, whereas São Paulo showed the lowest and least variable proportions.

As summarized in Figure 2F, the aggregated family refusal proportion for 2014–2024 was highest in Espírito Santo (49.86%), followed by Minas Gerais (43.99%), Rio de Janeiro (36.63%), and São Paulo (36.27%).

In Figure 3A, annual deceased-donor kidney transplant rates in Midwestern Brazil varied across the four federative units between 2014 and 2024. The Federal District generally presented the highest rates, although Goiás exceeded it between 2018 and 2020. Mato Grosso do Sul showed lower rates, while Mato Grosso recorded zero values in almost all years.

**Figure 3.**
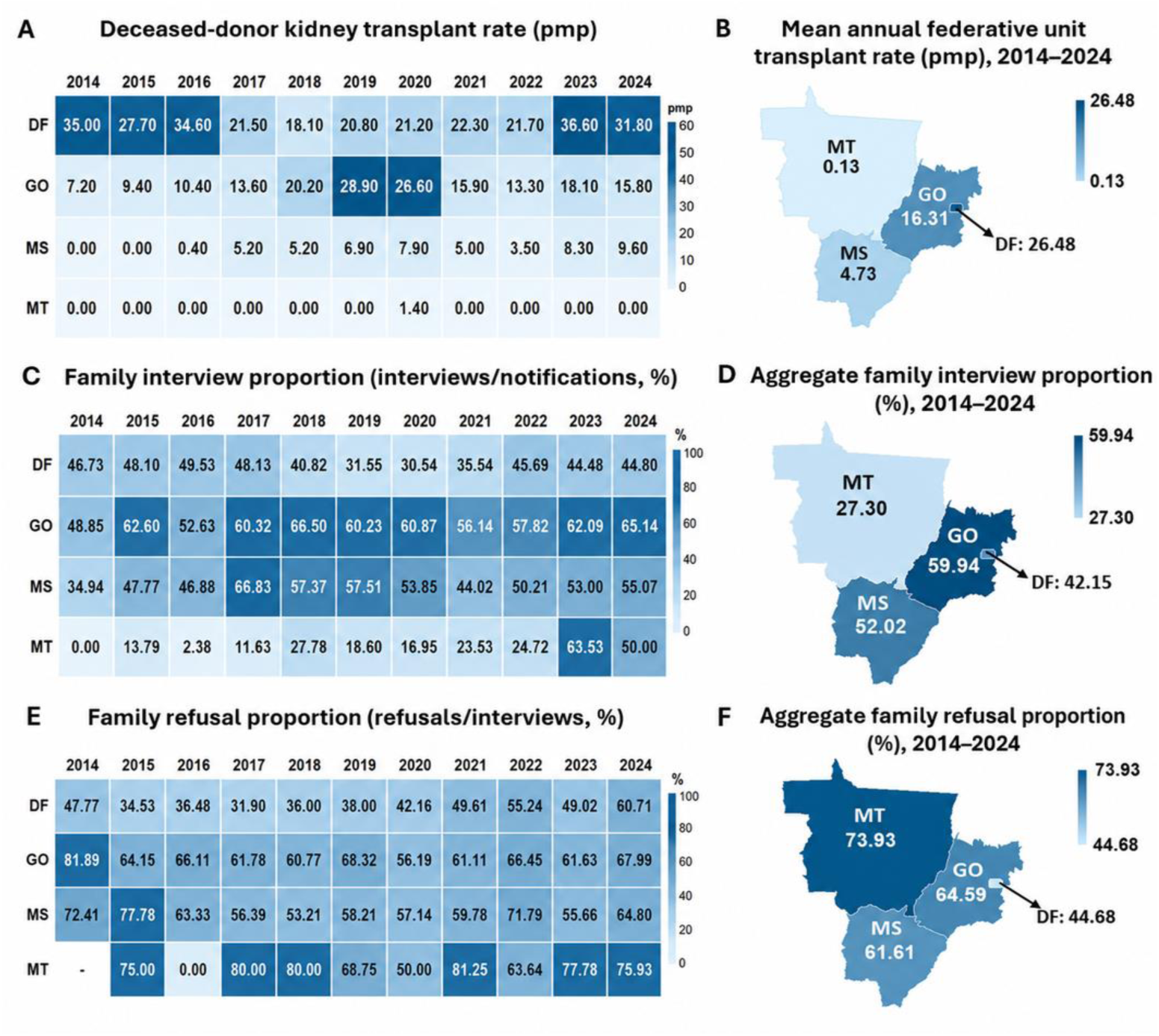
Deceased-donor kidney transplant activity and contextual indicators of the deceased-donation environment in Midwestern Brazil (Federal District [DF], Goiás [GO], Mato Grosso [MT], and Mato Grosso do Sul [MS]), 2014–2024. The deceased-donor kidney transplant rate per million population is shown as annual federative unit-level values (A) and as a geographic map of the mean annual transplant rate for 2014–2024 (B). Family interview proportion (interviews/notifications, %) is shown as annual federative unit-level values (C) and as a geographic map of the aggregated family interview proportion for 2014–2024, calculated as total interviews divided by total notifications over the study period (D). Family refusal proportion (refusals/interviews, %) is shown as annual federative unit-level values (E) and as a geographic map of the aggregated family refusal proportion for 2014–2024, calculated as total refusals divided by total interviews over the study period (F). Hyphens indicate years in which the proportion was not estimable because the denominator was equal to zero. True zero values were retained as 0.00 in the annual series. **Data source**: the Brazilian Transplant Registry (RBT/ABTO)¹⁰.

As shown in Figure 3B, the mean annual transplant rate for 2014–2024 was highest in the Federal District (26.48 pmp), followed by Goiás (16.31 pmp), Mato Grosso do Sul (4.73 pmp), and Mato Grosso (0.13 pmp).

In Figure 3C, family interview proportions also varied across the federative units. Goiás presented the highest aggregated proportion for 2014–2024 (59.94%), followed by Mato Grosso do Sul (52.02%), the Federal District (42.15%), and Mato Grosso (27.30%) (Figure 3D).

In Figure 3E, annual family refusal proportions were high and heterogeneous across the four federative units. As summarized in Figure 3F, the aggregated family refusal proportion for 2014–2024 was highest in Mato Grosso (73.93%), followed by Goiás (64.59%), Mato Grosso do Sul (61.61%), and the Federal District (44.68%).

In Figure 4A, annual deceased-donor kidney transplant rates in Northern Brazil were low and heterogeneous across the seven federative units between 2014 and 2024. Acre, Pará, Rondônia, and Amazonas showed fluctuating but generally low rates, whereas Amapá, Roraima, and Tocantins recorded 0.00 pmp throughout the study period.

**Figure 4.**
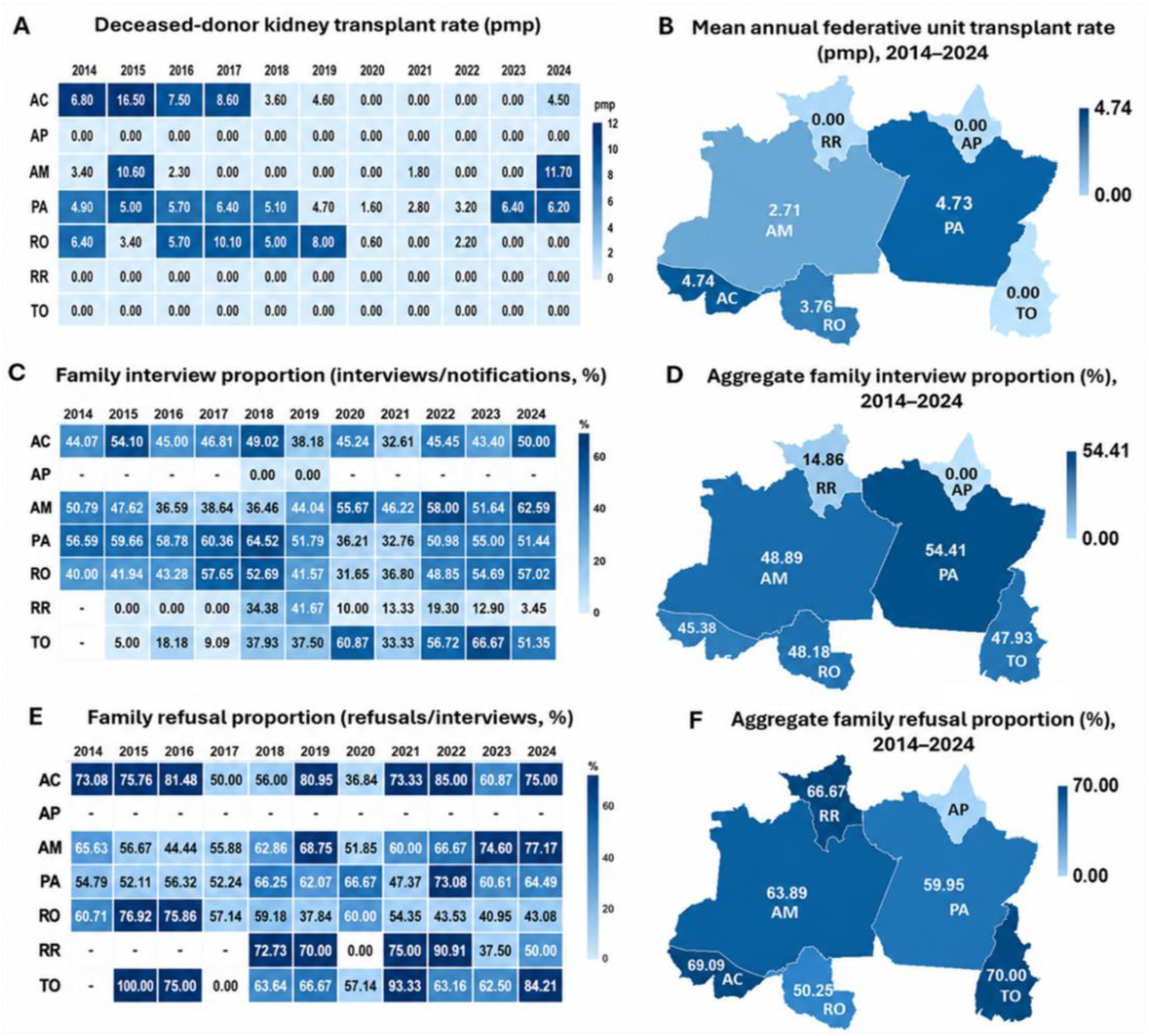
Deceased-donor kidney transplant activity and contextual indicators of the deceased-donation environment in Northern Brazil (Acre [AC], Amapá [AP], Amazonas [AM], Pará [PA], Rondônia [RO], Roraima [RR], and Tocantins [TO]), 2014–2024. The deceased-donor kidney transplant rate per million population is shown as annual federative unit-level values (A) and as a geographic map of the mean annual transplant rate for 2014–2024 (B). Family interview proportion (interviews/notifications, %) is shown as annual federative unit-level values (C) and as a geographic map of the aggregated family interview proportion for 2014–2024, calculated as total interviews divided by total notifications over the study period (D). Family refusal proportion (refusals/interviews, %) is shown as annual federative unit-level values (E) and as a geographic map of the aggregated family refusal proportion for 2014–2024, calculated as total refusals divided by total interviews over the study period (F). Hyphens indicate years in which the proportion was not estimable because the denominator was equal to zero. True zero values were retained as 0.00 in the annual series. **Data source**: the Brazilian Transplant Registry (RBT/ABTO)¹⁰.

As shown in Figure 4B, the highest mean annual transplant rates for 2014–2024 were observed in Acre (4.74 pmp) and Pará (4.73 pmp), followed by Rondônia (3.76 pmp) and Amazonas (2.71 pmp), while Amapá, Roraima, and Tocantins presented mean annual rates of 0.00 pmp.

In Figure 4C, family interview proportions also varied across the federative units, with Pará showing the highest aggregated proportion for 2014–2024 (54.41%), followed by Amazonas (48.89%), Rondônia (48.18%), Tocantins (47.93%), and Acre (45.38%). Lower values were observed in Roraima (14.86%), whereas Amapá presented 0.00% (Figure 4D). Some annual values were not estimable because the denominator was zero.

In Figure 4E, annual family refusal proportions were high in most federative units and showed substantial variation over time. As summarized in Figure 4F, the aggregated family refusal proportion for 2014–2024 was highest in Tocantins (70.00%), followed by Acre (69.09%), Roraima (66.67%), Amazonas (63.89%), Pará (59.95%), and Rondônia (50.25%), whereas no aggregated value was shown for Amapá.

In Figure 5A, deceased-donor kidney transplant rates in Northeastern Brazil showed marked heterogeneity across the nine federative units from 2014 to 2024. Pernambuco and Ceará consistently presented the highest rates, Rio Grande do Norte and Bahia showed intermediate values, and Piauí, Maranhão, Paraíba, and Alagoas had lower rates, while Sergipe recorded 0.00 pmp throughout the period.

**Figure 5.**
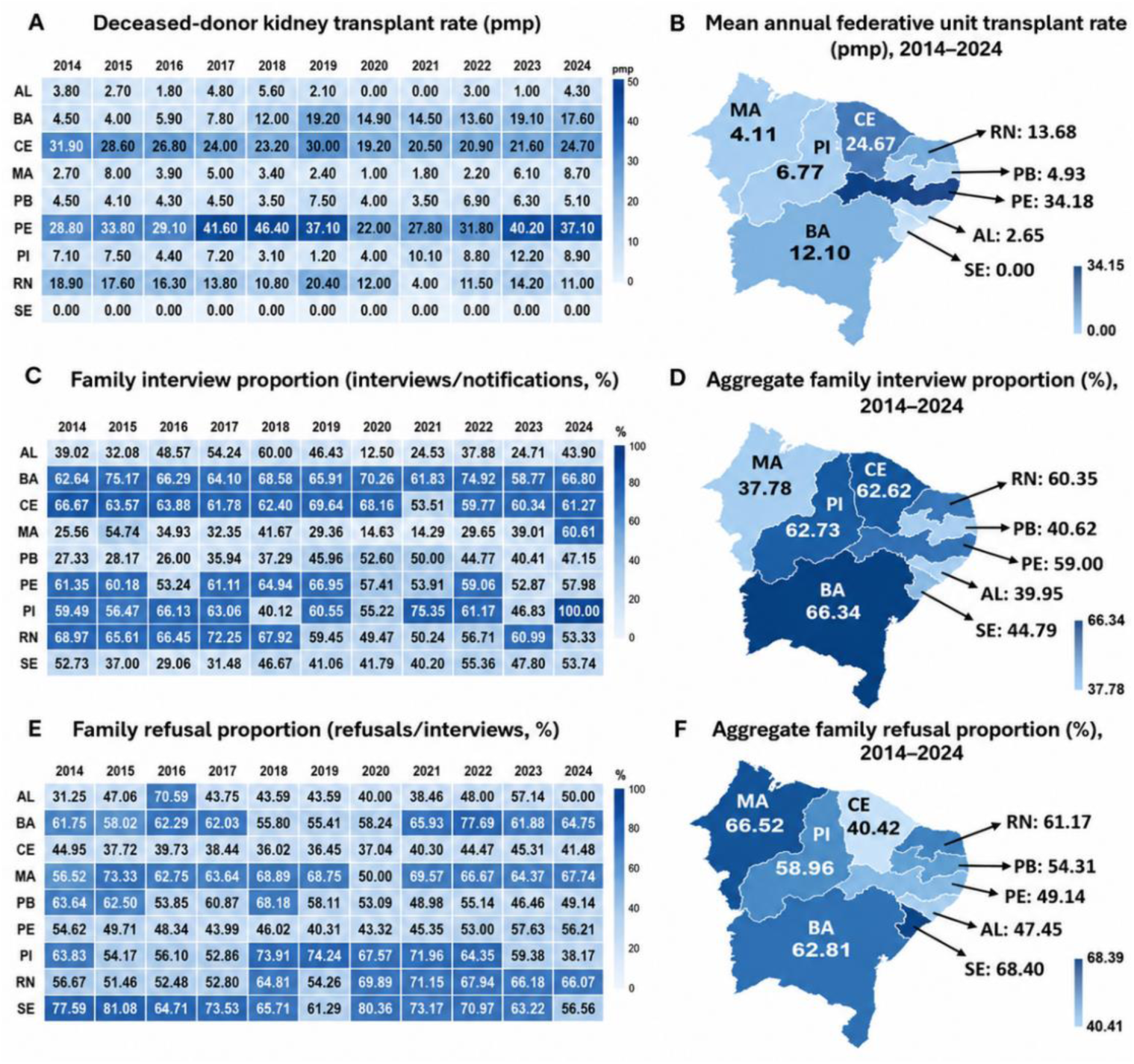
Deceased-donor kidney transplant activity and contextual indicators of the deceased-donation environment in Northeastern Brazil (Alagoas [AL], Bahia [BA], Ceará [CE], Maranhão [MA], Paraíba [PB], Pernambuco [PE], Piauí [PI], Rio Grande do Norte [RN], and Sergipe [SE]), 2014–2024. The deceased-donor kidney transplant rate per million population is shown as annual federative unit-level values (A) and as a geographic map of the mean annual transplant rate for 2014–2024 (B). Family interview proportion (interviews/notifications, %) is shown as annual federative unit-level values (C) and as a geographic map of the aggregated family interview proportion for 2014–2024, calculated as total interviews divided by total notifications over the study period (D). Family refusal proportion (refusals/interviews, %) is shown as annual federative unit-level values (E) and as a geographic map of the aggregated family refusal proportion for 2014–2024, calculated as total refusals divided by total interviews over the study period (F). **Data source**: the Brazilian Transplant Registry (RBT/ABTO)¹⁰.

As shown in Figure 5B, the highest mean annual transplant rates for 2014–2024 were observed in Pernambuco (34.18 pmp) and Ceará (24.67 pmp), followed by Rio Grande do Norte (13.68 pmp) and Bahia (12.10 pmp), whereas the remaining states presented lower values, including Sergipe (0.00 pmp).

In Figure 5C, family interview proportions also varied across the federative units. Bahia, Ceará, Rio Grande do Norte, and Pernambuco generally showed the highest proportions, whereas Sergipe, Alagoas, Paraíba, and Maranhão tended to present lower values; Piauí showed greater fluctuation.

In Figure 5D, the highest aggregated family interview proportions for 2014–2024 were observed in Bahia (66.34%), Piauí (62.73%), and Ceará (62.62%), while Maranhão presented the lowest value (37.78%).

In Figure 5E, family refusal proportions were high across the region, with Sergipe showing the highest values, followed by Maranhão, Bahia, and Rio Grande do Norte, whereas Ceará presented the lowest values overall. As summarized in Figure 5F, the aggregated family refusal proportion for 2014–2024 was highest in Sergipe (68.40%) and lowest in Ceará (40.42%).

### 3.4. Family refusal proportion across Brazilian macroregions and federative units: pooled 2014–2024 estimates and 2024 ranking

Table 3 presents, for each Brazilian macroregion, the federative unit with the highest aggregated family refusal proportion during 2014–2024, together with the corresponding aggregated family interview proportion. Both indicators are presented as denominator-weighted pooled period proportions. Tocantins presented the highest family refusal proportion in Northern Brazil (70.00%), with a family interview proportion of 47.93%; Mato Grosso, in Midwestern Brazil (73.93% and 27.30%); Sergipe, in Northeastern Brazil (68.40% and 44.79%); Espírito Santo, in Southeastern Brazil (49.86% and 53.73%); and Rio Grande do Sul, in Southern Brazil (43.19% and 66.53%).

**Table 3.** Federative units with the highest aggregated family refusal proportion in each Brazilian macroregion and the corresponding aggregated family interview proportion, Brazil, 2014–2024. **Data source:** the Brazilian Transplant Registry (RBT/ABTO)¹⁰. Abbreviations: UF, federative unit; TO, Tocantins; MT, Mato Grosso; SE, Sergipe; ES, Espírito Santo; RS, Rio Grande do Sul.

| Brazilian macroregions | UF | Family refusal<br>proportion (%) | Family interview<br>proportion (%) |
| --- | --- | --- | --- |
| North | TO | 70.00 | 47.93 |
| Midwest | MT | 73.93 | 27.30 |
| Northeast | SE | 68.40 | 44.79 |
| Southeast | ES | 49.86 | 53.73 |
| South | RS | 43.19 | 66.53 |

**Note:** Family refusal proportion was calculated as total family refusals divided by total family interviews over 2014–2024, multiplied by 100. Family interview proportion was calculated as total family interviews divided by total potential donor notifications over 2014–2024, multiplied by 100. These proportions correspond to denominator-weighted pooled period estimates rather than simple annual means.

Table 4 presents 2024 data for Brazilian federative units, ranked in descending order according to the proportion of family refusal. In 2024, the aggregate national family refusal proportion was 45.8%, rounded to 46.0%, corresponding to 4,083 refusals among 8,915 family interviews conducted in Brazil. The analysis by federative unit showed substantial heterogeneity, with rates ranging from 27.63% in Paraná to 84.21% in Tocantins. Amapá was not included in the ranking because no family interviews were recorded in 2024.

**Table 4.** Ranking of Brazilian federative units according to family refusal proportion in 2024. Data source: the Brazilian Transplant Registry (RBT/ABTO)¹⁰.

| Rank (2024) | UF | % | Rank (2024) | UF | % | Rank (2024) | UF | % |
| --- | --- | --- | --- | --- | --- | --- | --- | --- |
| 1st | TO | 84.21 | 10th | PA | 64.49 | 19th | RO | 43.08 |
| 2nd | AM | 77.17 | 11th | DF | 60.71 | 20th | MG | 41.97 |
| 3rd | MT | 75.93 | 12th | SE | 56.56 | 21st | CE | 41.48 |
| 4th | AC | 75.00 | 13th | PE | 56.21 | 22nd | SP | 39.74 |
| 5th | GO | 67.99 | 14th | AL | 50.00 | 23rd | PI | 36.79 |
| 6th | MA | 67.74 | 14th | RR | 50.00 | 24th | RJ | 34.97 |
| 7th | RN | 66.07 | 16th | PB | 49.14 | 25th | SC | 31.96 |
| 8th | MS | 64.80 | 17th | RS | 46.72 | 26th | PR | 27.63 |
| 9th | BA | 64.75 | 18th | ES | 44.90 | — | AP | — |

**Note:** Family refusal proportion was calculated as family refusals divided by family interviews in 2024, multiplied by 100. Amapá was not ranked because no family interviews were recorded in 2024.

The highest proportions of family refusal were observed in Tocantins (84.21%), Amazonas (77.17%), Mato Grosso (75.93%), and Acre (75.00%), all with rates equal to or above 70%. In the upper-intermediate range, between 60% and 69.9%, the following federative units stood out: Goiás (67.99%), Maranhão (67.74%), Rio Grande do Norte (66.07%), Mato Grosso do Sul (64.80%), Bahia (64.75%), Pará (64.49%), and the Federal District (60.71%). Conversely, the lowest rates were recorded in Paraná (27.63%), Santa Catarina (31.96%), Rio de Janeiro (34.97%), Piauí (36.79%), and São Paulo (39.74%).

## 4. Discussion

This nationwide descriptive study shows that deceased-donor kidney transplantation in Brazil increased between 2014 and 2024, but not at the same pace as the waiting list for kidney transplantation. While the annual number of deceased-donor kidney transplants increased by 25.88% and the deceased-donor kidney transplant rate per million population increased by 13.45%, the waiting list increased by 62.01%. Thus, the main system-level finding is not only the expansion of transplant activity, but also its occurrence alongside a larger increase in registered demand.

Hospitalization-based indicators also changed over the study period: the mean length of hospital stay decreased by 24.65%, and in-hospital mortality decreased by 18.13%. These concomitant reductions suggest a potentially favorable temporal pattern among SUS hospitalizations for deceased-donor kidney transplantation. However, because these indicators were derived from an administrative database and were not adjusted for patient-or center-level characteristics, they reflect patterns observed only within the subset of SUS hospitalizations captured in SIH/SUS.

At the macroregional level, the study identified heterogeneity in hospitalization-based indicators related to deceased-donor kidney transplantation. Shorter lengths of stay in Southeastern and Southern Brazil, longer stays in Midwestern and Northern Brazil, and variation in in-hospital mortality across macroregions illustrate territorial differences in these measures. Although the present analysis cannot identify the determinants of these differences, the findings reinforce that hospitalization-based patterns also varied territorially.

Beyond hospitalization-based measures, RBT/ABTO contextual indicators provided a complementary view of the deceased-donation environment. Potential donor notifications, family interviews, and family refusals represent key steps in the broader donation process, and their corresponding proportions summarize territorial variation in this pathway. Because these indicators are not kidney-specific, they were interpreted as contextual markers of the deceased-donation environment in which deceased-donor kidney transplantation occurs. Accordingly, deceased-donor kidney transplant activity was the only kidney-specific measure analyzed.

In descriptive terms, Southeastern and Southern Brazil concentrated higher transplant activity, with higher family interview proportions and lower family refusal proportions in several federative units. Northern and Midwestern Brazil showed lower transplant activity, lower family interview proportions, and higher family refusal proportions. The Northeast occupied an intermediate position in terms of transplant activity, with a smaller share than the Southeast and South but a larger share than the North and Midwest, while maintaining a high family refusal proportion.

Taken together, these findings describe heterogeneity in deceased-donor kidney transplantation in Brazil not only in transplant volume, but also in contextual indicators of the deceased-donation environment. Although the present study did not directly assess structural or organizational features, the regional patterns described here are compatible with prior descriptions of territorial variation in kidney transplantation in Brazil⁷.

Territorial variation in Brazil was observed not only between macroregions, but also within them. In Southeastern Brazil, São Paulo stood out with the highest mean annual transplant rate, the highest family interview proportion, and the lowest family refusal proportion. Espírito Santo showed the opposite profile, with the lowest transplant activity and the highest family refusal proportion, while Minas Gerais had the lowest family interview proportion. Rio de Janeiro and Minas Gerais occupied intermediate positions in transplant activity, with additional variation in family interview and refusal indicators.

Within-region heterogeneity was evident across all Brazilian macroregions. In Southern Brazil, all three federative units showed high mean annual transplant rates, with Rio Grande do Sul presenting the highest rate both regionally and nationally, while also showing the highest family refusal proportion within the Southern macroregion. Conversely, Santa Catarina and Paraná had lower refusal proportions, with Santa Catarina presenting the highest family interview proportion and Paraná the lowest family refusal proportion. These findings highlight substantial variation in donation-process indicators even among high-performing states.

In Northeastern Brazil, higher transplant activity was concentrated in Pernambuco and Ceará, whereas Sergipe recorded no local deceased-donor kidney transplant activity during the study period. Higher family interview proportions were observed in Bahia, Piauí, and Ceará, while Maranhão had the lowest family interview proportion. Family refusal proportions were highest in Sergipe, followed by Maranhão.

In Midwestern Brazil, the Federal District showed higher transplant activity and the lowest family refusal proportion, while Goiás had the highest family interview proportion. Mato Grosso do Sul occupied an intermediate position for transplant activity and contextual indicators, whereas Mato Grosso had the lowest mean annual transplant rate, the lowest family interview proportion, and the highest aggregated family refusal proportion in the region.

In Northern Brazil, Acre and Pará had the highest mean annual transplant rates, whereas Amapá, Roraima, and Tocantins did not record local transplant activity during the study period. Pará had the highest family interview proportion, Rondônia the lowest family refusal proportion, and Tocantins the highest aggregated family refusal proportion in the region. These within-region contrasts illustrate that macroregional summaries may fail to capture substantial heterogeneity at the federative-unit level.

Because organ procurement, allocation, transportation, and transplantation may occur across different federative units, comparisons between federative units should be interpreted with caution. Transplant activity recorded in a given federative unit does not necessarily correspond to kidneys originating from that same territory. The 2024 RBT/ABTO¹⁰ adds this interpretive layer by reporting the origin and destination of transplanted kidneys and describing real kidney utilization, which was 66% nationally; only the federative units of Ceará (58%) and Santa Catarina (57%) had utilization rates below 60%.

The flow matrix also shows that the federative units of MT, RO, SE, and TO did not perform local kidney transplantation in that dataset but contributed 186 kidneys to transplants performed elsewhere, while AC, AL, and PB transferred more than 70% of kidneys originating in their territories to other federative units. Conversely, kidneys from other federative units accounted for substantial shares of locally performed transplants in AM (64%), PE (40%), RS (38%), and DF (31%), and smaller shares in MG (18%) and SP (12%). These data reinforce that federative unit-level transplant indicators reflect an interconnected national system, rather than a one-to-one correspondence between local donation and local transplantation.

Within this same utilization context, approximately one-third of kidneys included in the 2024 RBT/ABTO analysis were not used for transplantation. This proportion should be interpreted cautiously, because the present study did not assess the reasons for kidney non-use and cannot distinguish clinical non-acceptance, donor-or organ-related factors, allocation issues, or logistical constraints related to organ non-use.

Nevertheless, logistics represents an important component of the donation and transplantation network. In this context, recent public policies have addressed logistical aspects of this network, including Law No. 14,858/2024, which establishes national priority for the transportation of organs, tissues, and human body parts intended for transplantation^11^.

Beyond these organ-flow and logistical considerations, the findings also show persistent regional and federative unit-level heterogeneity in kidney transplant rates per million population in 2024, further indicating an uneven distribution of transplant activity across the country. Among the contextual indicators examined, family refusal showed marked territorial variation. Although it represents only one component of the broader donation process, family authorization has been consistently described in the literature as a central step in deceased-donor organ donation programs^12–16^.

In the present study, family refusal proportions showed marked territorial heterogeneity across federative units and macroregions. In the aggregated 2014–2024 analysis by macroregion, the highest proportions were observed in Tocantins (North), Mato Grosso (Midwest), Sergipe (Northeast), Espírito Santo (Southeast), and Rio Grande do Sul (South). In 2024, the highest values were recorded in five federative units: Tocantins, Amazonas, Mato Grosso, Acre, and Goiás. From an interpretive standpoint, these findings suggest that donation-related heterogeneity is not fully captured by transplant counts alone and support the relevance of family refusal as a contextual marker of the deceased-donation environment.

In 2024, the aggregate national family refusal proportion in Brazil was 46%, highlighting the continued relevance of family refusal in deceased-donor organ donation. This finding is consistent with previous literature describing family refusal as a major barrier to the effectiveness of deceased-donor organ donation programs¹²⁻¹⁸. Although 67% of Brazilians reported being in favor of postmortem organ donation, many do not communicate this wish to their families, which is particularly relevant in a system that requires family consent¹⁹.

Frequently reported reasons for family refusal include fear of body mutilation, discomfort in discussing death, and limited knowledge about the donation process¹⁹. This evidence helps contextualize family refusal as a relevant marker for interpreting variation in deceased-donor kidney transplantation, as a single deceased donor may provide up to two transplantable kidneys. Thus, family refusal may result in the loss of more than one potential kidney transplant opportunity.

Against this background, recent public policies in Brazil have addressed dimensions related to awareness, professional training, and family support within the donation pathway. The National Policy for Awareness and Incentive for Organ and Tissue Donation and Transplantation, established by Law No. 14,722/2023, introduced guidelines for permanent educational campaigns, professional training, and family support²⁰. Within this policy context, the Electronic Authorization for Organ Donation (AEDO) represents another measure related to the formal registration of donation intention before the family approach process²¹.

From a policy perspective, these findings support the need for territorially tailored strategies to strengthen deceased-donation pathways, including public awareness campaigns, structured family approach protocols, professional training, logistical coordination, and continuous monitoring of donation-process indicators. Future studies using linked individual-level, donor-level, and center-level data are needed to better understand the determinants of regional variation and to evaluate the impact of recent policy initiatives on organ donation and transplantation outcomes.

## 5. Conclusion

In Brazil, deceased-donor kidney transplantation increased between 2014 and 2024; however, this expansion occurred alongside an even larger increase in the kidney transplant waiting list stock and persistent territorial heterogeneity. Differences across Brazilian macroregions and federative units extended beyond transplant activity, encompassing hospitalization-based indicators and contextual indicators of the deceased-donation environment. Potential donor notifications, family interviews, and family refusals represent distinct stages of the donation process and help describe territorial variation in the donation pathway. Thus, this study provides descriptive information that may support regional monitoring priorities, future analyses, and policy discussions.

## Study Limitations

The findings of this study should be interpreted in light of its limitations. Because this was a descriptive analysis based on aggregated data, it was not possible to examine individual-level clinical variables, account for differences in patient case mix or transplant center characteristics or draw inferences beyond the aggregate level. The parallel use of data sources with different structures, scopes, and levels of aggregation also requires caution, as indicators derived from one database do not necessarily reflect the same underlying population or denominator as those derived from another. The results may also have been affected by data completeness, reporting quality, internal inconsistencies in registry records, and potential misclassification.

## Authors’ Contributions

Conceptualization: MBC, FTB; Data curation: MBC; Formal analysis: MBC; Investigation: MBC; Methodology: MBC, FTB; Project administration: MBC, FTB; Resources: MBC, FTB; Software: MBC; Supervision: FTB; Validation: MBC, FTB; Visualization: MBC, FTB; Writing – original draft: MBC; Writing – review and editing: MBC, FTB. All authors have approved the final version of the manuscript.

## Data Availability

The data analyzed in this study are publicly available from DATASUS/SIH-SUS and the Brazilian Transplant Registry (RBT/ABTO), and Brazilian National Transplant System (SNT) as cited in the Methods.

